# Artificial intelligence analytics applied to body mass index global burden of disease worldwide cohort data derives a multiple regression formula with population attributable fraction risk factor coefficients testable by all nine Bradford Hill causality criteria

**DOI:** 10.1101/2020.07.27.20162487

**Authors:** David K Cundiff, Chunyi Wu

**Affiliations:** Long Beach, California, USA; Area Specialist Lead in Epidemiology and Statistics, Michigan Medicine, Ann Arbor, Michigan, USA; Volunteer collaborators with the Institute of Health Metrics and Evaluation, Seattle, Washington, USA

## Abstract

**Background:** Artificial intelligence (AI) analytics have not been applied to global burden of disease (GBD) risk factor data to study population health. The comparative risk assessment (CRA) systematic literature review-based methodology for population attributable fractions (PAFs in percent’s) calculations has not been utilised for quantifying dietary and other risk factors for body mass index kg/M^2^ (BMI).

**Methods:** Institute of Health Metrics and Evaluation (IHME) staff and volunteer collaborators analysed over 12,000 GBD risk factor surveys of people from 195 countries and synthesized the data into representative mean cohort BMI and risk factor values. We formatted IHME GBD data relevant to BMI and associated risk factors. We empirically explored the univariate and multiple regression correlations of BMI risk factors with worldwide BMI to derive a BMI multiple regression formula (BMI formula). Main outcome measures included the performances of the BMI formula when tested with all nine Bradford Hill causality criteria each scored on a 0-5 scale: 0=negative to 5=very strong support.

**Findings:** The BMI formula derived, with all foods in kilocalories/day (kcal/day), BMI formula risk factor coefficients were adjusted to equate with their PAFs. BMI increasing foods had “+” signs and BMI decreasing foods “-” signs. Total BMI formula PAF=80.96%. BMI formula=(0.37%*processed meat + 4.23%*red meat + 0.02%*fish + 2.24%*milk + 5.67%*poultry + 1.77%*eggs + 0.34%*alcohol + 0.99%*sugary beverages + 0.04%*corn + 0.72%*potatoes + 8.48%*saturated fatty acids + 3.89%*polyunsaturated fatty acids + 0.27%*trans fatty acids - 2.99%*fruit - 4.07%*vegetables - 0.37%*nuts and seeds - 0.45%*whole grains - 1.49%*legumes - 8.62%*rice - 0.10%*sweet potatoes - 7.45% physical activity (METs/week) - 20.38%*child underweight + 6.02%*sex (male=1, female=2))*0.05012 + 21.77. BMI formula versus BMI: r=0.907, 95% CI: 0.903 to 0.911, *p*<0.0001. Bradford Hill causality criteria test scores (0-5): (1) strength=5, (2) experimentation=5, (3) consistency=5, (4) dose-response=5, (5) temporality=5, (6) analogy=4, (7), plausibility=5, (8) specificity=5, and (9) coherence=5. Total score=44/45.

**Interpretation:** Nine Bradford Hill causality criteria strongly supported a causal relationship between the BMI formula derived and mean BMIs of worldwide cohorts. The artificial intelligence methodology introduced could inform individual, clinical, and public health strategies regarding overweight/obesity prevention/treatment and other health outcomes.

**Funding:** None

**Research in context:** *Evidence before this study:* Comparative risk assessment (CRA) systematic literature review-based methodology has been used in worldwide global burden of disease (GBD) analysis to determine population attributable fraction(s) (PAF(s)) for one or more risk factors for various health outcomes. So far, CRA has not been applied to derive PAFs for dietary and other risk factors for worldwide BMI. Artificial intelligence (AI) analytics has not yet been applied to worldwide GBD data as an alternative to the CRA methodology for determining risk factor PAFs for health outcomes.

*Added value of this study□:* A multiple regression derived BMI formula (BMI formula) including PAFs of 20 dietary risk factors, physical activity, childhood severe underweight, and sex satisfied all nine Bradford Hill causality criteria. The BMI formula also plausibly predicted the long-term BMI outcomes related to various dietary and physical activity scenarios. All the BMI formula’s 24 risk factor PAFs were consistent in sign (+ or -) with the preponderance of previously published studies on those risk factors related to BMI.

*Implications of all the available evidence:* The AI analytics methodology of GBD data modeling of BMI and associated risk factors infers causality of the BMI formula estimates with BMI worldwide and BMIs of subsets. This methodology may enable multiple regression formulas for risk factors of health outcomes for a range of non-communicable diseases—testable by Bradford Hill causality criteria.

## Introduction

Comparable risk assessment (CRA) methodology by the World Health Organization and the Institute of Health Metrics and Evaluation (IHME) is defined as, “the systematic evaluation of the changes in population health which result from modifying the population distribution of exposure to a risk factor or a group of risk factors.”^1, 2^ While CRA methodologies have used body mass index kg/M^2^ (BMI) in risk factor—health outcome pairs for numerous health outcomes (e.g., BMI-ischemic heart disease, BMI-colon cancer, BMI-type 2 diabetes, etc.), CRA has not been attempted to quantify the largely nutritional risk factors related to BMI itself.

Influential Stanford University meta researcher, Dr. John Ioannidis provided a likely reason for this in an editorial in the *BMJ* titled, “Implausible results in human nutrition research.” He said, “definitive solutions won’t come from another million observational papers or small randomized trials.”^**3**^ Dr. Ioannidis called for radical reform of nutritional epidemiology methodologies used to influence food/agricultural policies and to produce dietary guidelines for clinicians and the public.^**4**^

According to IBM Cloud Education, “At its simplest form, artificial intelligence (AI) is a field, which combines computer science and robust datasets, to enable problem-solving.”^5^ AI will be used instead of the CRA methodology in this paper.

Satisfying Bradford Hill causality criteria is considered validating in epidemiological research.^6^ We used Bradford Hill causality criteria to test the BMI formula. We hypothesized that AI analytics with GBD big data would be able to quantify the determinants of worldwide cohort BMIs and thereby to better our understanding the dietary and other risk factors driving the obesity epidemic. We hope this paper answers Dr. Ioannidis’ call for innovation in epidemiology, particularly nutritional epidemiology.

## Methods

As volunteer collaborators with the IHME we received raw GBD ecological data (≈1.4 Gigabytes) on mean BMIs of male and female cohorts 15-49 years old and 50-69 years old from each year 1990-2017 from 195 countries and 365 subnational locations (n=1120 cohorts). We also utilised GBD data on exposures to 32 risk factors and covariates potentially related to BMI.

Food risk factors came from surveys of individuals as g/day. IHME dietary covariate data originally came from Food and Agriculture Organisation data on animal and plant food commodities available percapita in countries worldwide—as opposed to consumption data from participant interviews.^7^ Supplementary Table 1 lists the relevant GBD risk factors, covariates, and other available variables with definitions of those risk factor exposures.^8^

GBD worldwide citations of over 12,000 surveys constituting ecological data inputs for this analysis are available from IHME.^7^ The main characteristics of IHME GBD data sources for BMI, the protocol for the GBD study, and all risk factor values have been published by IHME GBD data researchers and discussed elsewhere.^9-12^ These included detailed descriptions of categories of input data, potentially important biases, and methodologies of analysis. We did not clean or pre-process any of the GBD data. GBD cohort risk factor and BMI data from the IHME had no missing records. The updated 2019 raw data with variables we used for this analysis may be obtained by volunteer researchers collaborating with IHME.^13^

To maximally utilise the available data, we averaged the values for ages 15-49 years old together with 50-69 years old for BMI and for each risk factor exposure for each male and female cohort for each year. Finally, for each male and female cohort, data from all 28 years (1990-2017) on mean BMI and on each of the risk factor exposures were averaged using the computer software program R.

To weigh the country and subnational data according to population, internet searches (mostly Wikipedia) yielded the most recent population estimates for countries and subnational states, provinces, and regions. World population data from the World Bank and the Organisation for Economic Co-operation and Development could not be used because they did not include all 195 countries or any subnational data.

Using the above-described formatted dataset of risk factors, covariates, and BMIs, a software program in R generated a population-weighted analysis dataset. Each male or female cohort in the population-weighted analysis dataset represented approximately 1 million people (range: < 100,000 to 1.5 million). The analysis dataset had n=7846 cohorts (rows of data), half male and half female, representative of over seven billion people.

Supplementary Table 2 details how we converted omega-3 fatty acid g/day to fish g/day using data on the omega-3 fatty acid content of frequently eaten fish from the National Institutes of Health Office of Dietary Supplements (USA).^14^ As shown in Supplementary Table 3, we converted all of the animal and plant food data, including alcohol and sugary beverage consumption, from g/day to kcal/day. For the g to kcal conversions, we used the Nutritionix track app,^15^ which tracks types and quantities of foods consumed. Saturated fatty acids (SFA) risk factor (0-1 portion of the entire diet) was not available with GBD data from 2017, so GBD SFA risk factor data from 2016 was used. Polyunsaturated fatty acid (PUFA) and trans fatty acid (TFA) GBD risk factor data from 2017 (0-1 portion of the entire diet) were also utilised. These fatty acid data were converted to kcal/day by multiplying the fatty acid 0-1 portion of the entire diet by the kcal/day available for each cohort.

### Statistical methods

To determine the strengths of the risk factor correlations with mean BMIs of population weighted worldwide cohorts (7846 cohorts) or subgroups of cohorts (e.g., continents, socio-demographic quartiles, etc.), we utilised Pearson correlation coefficients: r, 95% confidence intervals (95% CIs), and *p* values.

In this first attempt ever to use AI analytics with GBD data, we determined our methodology for deriving a BMI formula as we proceeded by experimenting with strategies to optimise the functioning of candidate BMI formulas. We

1. included as many as possible of the available dietary variables,
2. combined dietary variables if appropriate, and
3. included physical activity and other plausibly informative variables.

Appendix 1 explains the use of Bradford Hill causality criteria to assess whether the risk factors in the BMI formula derived accurately modeled the worldwide mean BMI. Briefly, we tested the BMI formula output with the Bradford Hill causality criteria (1) strength, (2) experimentation, (3) consistency, (4) dose response, (5) temporality, (6 analogy, (7) plausibility, (8) specificity, and (9) coherence. For each criterion, we used a 0-5 scale to assess the magnitude of support of the BMI formula output being causally related to the BMIs worldwide (0=no support of causality to 5=very strong support of causality, total possible points=45.

In determining the variables to include and exclude in worldwide BMI formula, we set the statistical threshold for a variable to enter and to remain in the formula at *p*<0.25. We used SAS and SAS Studio statistical software 9.4 (SAS Institute, Cary, NC) for the data analysis.

## Results

Table 1 shows the basic statistics and univariate correlations of mean cohort BMI worldwide with mean dietary and other risk factor values. Whole grains, legumes, rice, and sweet potatoes negatively correlated with BMI. We designated them, “BMI decreasing foods.” The six animal foods (processed meat, red meat, fish, milk, poultry, and eggs), alcohol, sugary beverages, corn availability, potato availability, added SFA, added PUFA, and added TFA all positively correlated with BMI. We designated them, “BMI increasing foods.”

**Table 1.** BMI risk factor basic statistics□ (n=7846)

| BMI versus BMI risk factors worldwide n=7846<br>□ | Mean | SD | Min | Max | r | 95%<br>CI low | 95%<br>CI high | p | R<sup>2</sup> |
| --- | --- | --- | --- | --- | --- | --- | --- | --- | --- |
| BMI kg/M<sup>2</sup> | 21.77 | 2.290 | 17.95 | 29.39 |  |  |  |  |  |
| Processed meat Kcal/day | 5.334 | 9.720 | 0.20 | 68.77 | 0.594 | 0.579 | 0.608 | <.0001 | 0.3527 |
| Red meat Kcal/day | 50.27 | 45.13 | 3.21 | 235.95 | 0.656 | 0.643 | 0.668 | <.0001 | 0.4304 |
| Fish Kcal/day | 9.985 | 36.52 | 0.40 | 370.36 | 0.102 | 0.080 | 0.124 | <.0001 | 0.0104 |
| Milk Kcal/day | 25.04 | 27.05 | 1.06 | 146.8 | 0.676 | 0.664 | 0.688 | <.0001 | 0.4573 |
| Poultry Kcal/day available | 44.32 | 50.08 | 1.06 | 411.9 | 0.809 | 0.801 | 0.816 | <.0001 | 0.6539 |
| Eggs Kcal/day available | 19.36 | 14.71 | 0.79 | 69.64 | 0.684 | 0.672 | 0.695 | <.0001 | 0.4672 |
| Alcohol Kcal/day | 81.03 | 57.33 | 4.25 | 429.8 | 0.146 | 0.124 | 0.168 | <.0001 | 0.0213 |
| Sugary beverages Kcal/day | 298.4 | 152.4 | 72.91 | 1472 | 0.131 | 0.109 | 0.152 | <.0001 | 0.0170 |
| Corn Kcal/day available | 34.72 | 48.28 | 0.16 | 305.2 | 0.076 | 0.054 | 0.098 | <.0001 | 0.0058 |
| Potatoes Kcal/day available | 84.04 | 74.60 | 3.07 | 533.9 | 0.210 | 0.188 | 0.231 | <.0001 | 0.0439 |
| Saturated fatty acids Kcal/day | 190.6 | 63.94 | 70.79 | 481.1 | 0.703 | 0.692 | 0.714 | <.0001 | 0.4946 |
| PUFAs Kcal/day | 81.10 | 73.44 | 2.93 | 381.3 | 0.730 | 0.720 | 0.740 | <.0001 | 0.5330 |
| Trans fatty acids Kcal/day | 13.23 | 13.65 | 1.99 | 77.76 | 0.475 | 0.458 | 0.492 | <.0001 | 0.2256 |
| Fruits Kcal/day | 40.21 | 22.50 | 3.58 | 161.4 | 0.616 | 0.602 | 0.630 | <.0001 | 0.3798 |
| Vegetables Kcal/day | 79.76 | 43.12 | 9.48 | 304.2 | 0.511 | 0.495 | 0.527 | <.0001 | 0.2612 |
| Nuts and seeds Kcal/day | 8.414 | 8.357 | 0.05 | 103.0 | 0.474 | 0.457 | 0.491 | <.0001 | 0.2248 |
| Whole grains Kcal/day | 55.65 | 30.93 | 1.14 | 235.1 | -0.203 | -0.224 | -0.182 | <.0001 | 0.0412 |
| Legumes Kcal/day | 51.74 | 32.23 | 0.51 | 194.7 | -0.384 | -0.402 | -0.364 | <.0001 | 0.1471 |
| Rice Kcal/day available | 141.9 | 116.3 | 1.42 | 461.8 | -0.557 | -0.572 | -0.542 | <.0001 | 0.3104 |
| Sweet potatoes Kcal/day available | 22.76 | 35.95 | 0.02 | 364.7 | -0.148 | -0.169 | -0.126 | <.0001 | 0.0218 |
| Physical activity METs | 4714 | 1368 | 1609 | 7669 | -0.443 | -0.461 | -0.425 | <.0001 | 0.1964 |
| Child underweight by >2SD | 0.1862 | 0.1707 | 0.0039 | 0.5300 | -0.794 | -0.802 | -0.786 | <.0001 | 0.6306 |
| Sex male 1 and female 2 | 1.50 | 0.500 | 1.00 | 2.00 | 0.124 | 0.102 | 0.145 | <.0001 | 0.0153 |
| Total Kcal/day available | 2574 | 418 | 1579 | 3898 | 0.840 | 0.834 | 0.847 | <.0001 | 0.7059 |
| Stop breast feeding <6 months | 0.1193 | 0.0555 | 0.0159 | 0.2400 | 0.799 | 0.791 | 0.807 | <.0001 | 0.6383 |
| Sodium g/d | 4.45 | 2.34 | 1.33 | 9.21 | -0.017 | -0.039 | 0.005 | 0.1268 | 0.0000 |
| Calcium g/d | 0.301 | 0.179 | 0.081 | 1.04 | 0.765 | 0.756 | 0.774 | <.0001 | 0.5850 |
| Dietary fiber g/d | 9.21 | 3.15 | 2.72 | 22.68 | 0.309 | 0.289 | 0.329 | <.0001 | 0.0960 |
| Fasting plasma glucose mmol/L | 4.30 | 0.350 | 3.32 | 5.58 | 0.557 | 0.542 | 0.572 | <.0001 | 0.3110 |
| LDL cholesterol mmol/L | 2.35 | 0.400 | 1.27 | 3.25 | 0.756 | 0.746 | 0.765 | <.0001 | 0.5710 |
| Systolic BP mm Hg | 133.9 | 4.320 | 123.4 | 147.9 | 0.102 | 0.080 | 0.124 | <.0001 | 0.0100 |
| Socio-demographic index (0-1) | 0.543 | 0.174 | 0.112 | 0.900 | 0.733 | 0.722 | 0.743 | <.0001 | 0.5370 |

□ See Supplementary Table 1 for definitions

In Supplementary Table 4, which shows BMI risk factor to risk factor correlations, corn availability (kcal/day percapita, a covariate) correlated moderately strongly with sugary beverages (r=0.419, 95% CI 0.400 to 0.437, *p*<0.0001), suggesting that high fructose corn syrup may have accounted for the unexpected positive correlation of corn with BMI.

The strong positive correlations of fruits, vegetables, and nuts and seeds with BMI (Table 1) were unexpected. These findings suggested likely multicollinearity of fruits, vegetables, and nuts and seeds with BMI increasing foods (when an independent variable is highly correlated with one or more known or unknown other independent variables). Supplementary Table 4 shows that fruits, vegetables and nuts and seeds strongly positively correlated with 10 of the 13 dietary variables that positively correlated with BMI (six animal-based foods, alcohol, SFA, PUFA, and TFA). Fruits, vegetables and nuts and seeds negatively correlated with the four other plant-based foods (whole grains, legumes, rice, and sweet potatoes), except fruits was not significantly correlated with whole grains. Together, these findings strongly suggested that multicollinearities accounted for the positive correlations of fruits, vegetables, and nuts and seeds with BMI.

Supplementary Table 5 shows the Excel spreadsheet calculations, which were coordinated with SAS multiple regressions involved in the derivation of the worldwide BMI formula. Appendix 2 detailed the steps in the multiple regression formula derivation of BMI (dependent variable) versus BMI risk factors (independent variables). As seen in Supplementary Table 5 and described in Appendix 2, the BMI formula versus BMI variance (R^2^=0.8096) * 100 determined the total PAF (80.96%) accounted for by the BMI formula. The 23 risk factor coefficients could then be equated to the PAFs of those individual risk factors. The resulting BMI formula is shown below (all foods in kcal/day):

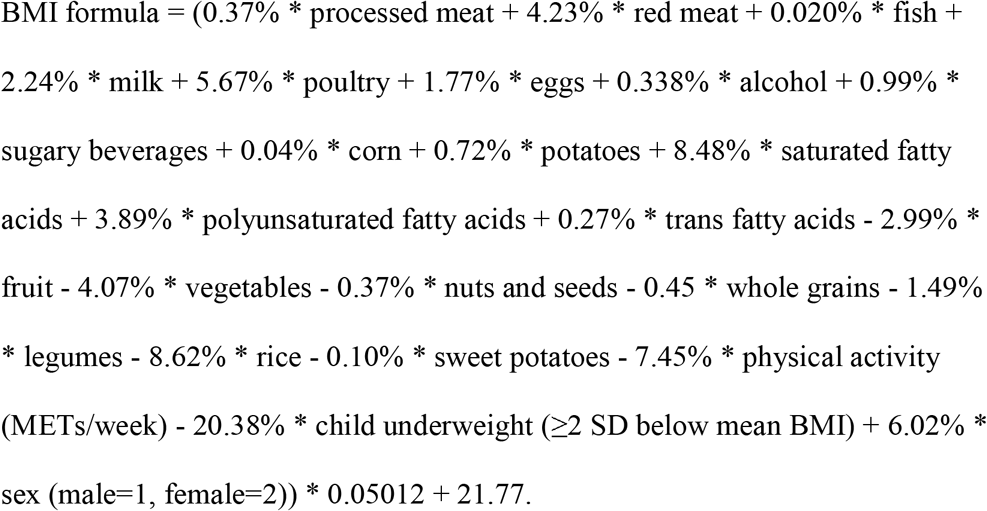

### BMI formula output analysed by Bradford Hill causality criteria

As mentioned in the methods and detailed in Appendix 1, the nine Bradford Hill causality criteria tested the functionality of the BMI formula:

1. Strength score=5 for r>0.500, *p*<0.0001—The BMI formula’s output regressed with mean BMI of worldwide cohorts gave r= 0.907 (95% CI: 0.903 to 0.911) *p*<0.0001), R^2^=0.8096, total percent weight=80.96%.
2. Experiment score=5 for all 20 bootstrap trial BMI formulas with r>0.500, *p*<0.0001)— Table 2 shows bootstrapping the BMI formula related to mean worldwide BMI with 20 trials (repeated sampling from the worldwide analysis dataset with replacements^16^). Each trial had 100 randomly selected cohorts to generate a unique BMI formula. As Table 2 shows, the mean values for BMI and the risk factor PAFs were all quite close to the mean values for BMI and BMI risk factors in the worldwide BMI formula (last column).

**Table 2.** Bootstrap validation experiment: 20 BMI formulas each with 100 random cohorts.
3. Consistency score=5 for the mean of absolute differences of BMI formula outputs and mean BMI < 0.300 kg/M^2^—In Table 3, we used 37 subsets of worldwide BMI and risk factor data to test the consistency of BMI formula outputs, utilising the 20 bootstrap trials (#2 Experiment) to generate 80% confidence intervals for each subset. Table 3 shows the average absolute difference between the 37 subgroups mean BMIs and BMI formula outputs averaged 0.252 kg/M^2^.

| Worldwide BMI and BMI risk factors | RCT1 | RCT2 | RCT3 | RCT4 | RCT5 | RCT6 | RCT7 | RCT8 | RCT9 | RCT 10 | RCT 11 | RCT 12 | RCT 13 |
| --- | --- | --- | --- | --- | --- | --- | --- | --- | --- | --- | --- | --- | --- |
| BMI kg/M<sup>2</sup> | 21.71 | 22.04 | 21.68 | 21.92 | 21.57 | 21.42 | 21.79 | 22.18 | 21.75 | 21.85 | 21.49 | 21.88 | 21.71 |
| Processed meat | 0.33 | 0.32 | 0.34 | 0.34 | 0.28 | 0.31 | 0.32 | 0.39 | 0.57 | 0.36 | 0.21 | 0.26 | 0.28 |
| Red meat | 3.95 | 3.73 | 4.92 | 3.47 | 3.80 | 3.70 | 3.45 | 4.89 | 5.61 | 3.69 | 3.56 | 3.20 | 3.16 |
| Fish | 0.03 | 0.02 | 0.03 | 0.01 | 0.64 | 0.03 | 0.03 | 0.03 | 0.04 | 0.02 | 0.02 | 0.03 | 0.49 |
| Milk | 2.13 | 2.29 | 2.40 | 2.19 | 2.04 | 1.95 | 1.89 | 2.39 | 2.66 | 2.14 | 1.68 | 1.83 | 1.43 |
| Poultry | 6.62 | 6.24 | 6.60 | 4.82 | 5.36 | 4.33 | 5.92 | 6.72 | 5.41 | 4.84 | 4.48 | 6.55 | 4.15 |
| Eggs | 1.81 | 1.68 | 1.84 | 1.45 | 2.17 | 1.70 | 1.57 | 1.85 | 2.10 | 1.65 | 1.59 | 1.46 | 1.39 |
| Alcohol | 0.09 | 0.74 | 0.03 | 0.28 | 0.05 | 0.04 | 0.77 | 0.59 | 1.65 | 0.38 | 0.04 | 0.00 | 0.01 |
| Sugary beverages | 1.90 | 2.25 | 6.36 | 0.78 | 4.79 | 2.44 | 0.43 | 3.96 | 0.30 | 1.55 | 2.04 | 3.43 | 2.90 |
| Corn | 0.04 | 0.00 | 0.09 | 0.01 | 0.26 | 0.02 | 0.14 | 0.27 | 0.00 | 0.01 | 0.32 | 0.28 | 0.32 |
| Potatoes | 0.23 | 0.10 | 0.60 | 0.67 | 0.69 | 0.15 | 0.11 | 1.77 | 1.26 | 0.68 | 1.39 | 0.02 | 1.34 |
| SFA | 8.56 | 7.14 | 8.55 | 6.96 | 9.12 | 9.81 | 7.44 | 8.52 | 8.80 | 7.75 | 6.66 | 7.10 | 5.97 |
| PUFA | 4.30 | 3.97 | 4.31 | 3.16 | 4.09 | 3.59 | 4.09 | 4.12 | 3.96 | 3.66 | 2.84 | 3.30 | 2.84 |
| TFA | 0.33 | 0.29 | 0.23 | 0.23 | 0.26 | 0.14 | 0.29 | 0.30 | 0.38 | 0.26 | 0.19 | 0.35 | 0.14 |
| Fruits | 3.18 | 2.55 | 3.99 | 3.25 | 3.88 | 2.39 | 1.95 | 3.38 | 2.44 | 2.00 | 2.31 | 2.82 | 2.65 |
| Vegetables | 3.22 | 3.39 | 4.70 | 5.83 | 3.67 | 2.95 | 4.38 | 3.79 | 5.06 | 4.04 | 5.51 | 4.46 | 1.87 |
| Nuts and seeds | 0.22 | 0.32 | 0.51 | 0.30 | 0.55 | 0.24 | 0.36 | 0.59 | 0.60 | 0.25 | 0.07 | 0.13 | 0.13 |
| Whole grains | 0.14 | 0.40 | 0.24 | 0.85 | 0.26 | 0.68 | 0.18 | 0.01 | 0.12 | 0.29 | 1.32 | 0.53 | 0.17 |
| Legumes | 2.86 | 1.41 | 0.71 | 1.38 | 1.44 | 1.82 | 1.44 | 1.06 | 1.65 | 1.48 | 1.12 | 1.71 | 1.33 |
| Rice | 7.24 | 6.91 | 7.76 | 9.80 | 10.26 | 8.25 | 6.35 | 9.19 | 5.84 | 4.31 | 12.17 | 8.20 | 7.82 |
| Sweet potatoes | 0.05 | 0.29 | 0.12 | 0.20 | 0.04 | 0.07 | 0.22 | 0.12 | 0.17 | 0.17 | 0.02 | 0.30 | 0.05 |
| Physical activity | 6.65 | 10.11 | 3.69 | 9.29 | 0.66 | 5.71 | 14.65 | 2.63 | 3.81 | 10.17 | 7.80 | 7.37 | 6.38 |
| Child underweight | 8.86 | 8.41 | 5.17 | 5.47 | 9.68 | 21.13 | 23.33 | 15.63 | 20.69 | 27.27 | 22.28 | 15.50 | 23.20 |
| Sex M=1, F=2 | 21.20 | 19.02 | 13.78 | 18.92 | 15.67 | 9.49 | 1.65 | 7.23 | 5.06 | 4.36 | 4.29 | 10.32 | 9.07 |
| Total % weights | 83.94 | 81.57 | 76.96 | 79.67 | 79.66 | 80.95 | 80.97 | 79.45 | 78.17 | 81.35 | 81.91 | 79.14 | 77.11 |

| Worldwide BMI and BMI risk factors | RCT 14 | RCT 15 | RCT 16 | RCT 17 | RCT 18 | RCT 19 | RCT 20 | Mean RCTs 1-20 | SD | 80% CI low | 80% CI high | World mean n=7846 |
| --- | --- | --- | --- | --- | --- | --- | --- | --- | --- | --- | --- | --- |
| BMI kg/M2 | 21.79 | 21.65 | 21.81 | 21.81 | 22.01 | 21.76 | 21.79 | 21.78 | 0.13 | 21.49 | 22.04 | 21.77 |
| Processed meat | 0.30 | 0.53 | 0.37 | 0.41 | 0.28 | 0.31 | 0.55 | 0.36 | 0.09 | 0.26 | 0.55 | 0.37 |
| Red meat | 2.65 | 4.77 | 3.57 | 4.55 | 2.47 | 3.49 | 5.71 | 3.95 | 0.84 | 2.65 | 5.61 | 4.23 |
| Fish | 0.02 | 0.02 | 0.01 | 0.03 | 0.02 | 0.56 | 0.70 | 0.12 | 0.23 | 0.01 | 0.64 | 0.02 |
| Milk | 1.84 | 2.29 | 1.79 | 2.51 | 1.58 | 1.80 | 2.50 | 2.10 | 0.33 | 1.58 | 2.51 | 2.24 |
| Poultry | 4.55 | 4.91 | 4.87 | 5.66 | 4.66 | 4.52 | 6.87 | 5.43 | 0.87 | 4.33 | 6.72 | 5.67 |
| Eggs | 1.47 | 1.51 | 1.33 | 2.01 | 1.32 | 1.61 | 2.08 | 1.69 | 0.25 | 1.33 | 2.10 | 1.77 |
| Alcohol | 0.01 | 2.10 | 0.66 | 0.67 | 0.01 | 0.19 | 1.32 | 0.46 | 0.56 | 0.01 | 1.65 | 0.34 |
| Sugary beverages | 2.58 | 1.31 | 0.79 | 1.11 | 0.99 | 0.61 | 0.29 | 1.90 | 1.55 | 0.30 | 4.79 | 0.99 |
| Corn | 0.37 | 0.00 | 0.54 | 0.11 | 0.22 | 0.35 | 0.00 | 0.15 | 0.16 | 0.00 | 0.37 | 0.04 |
| Potatoes | 0.49 | 0.06 | 1.49 | 0.26 | 0.16 | 1.71 | 0.21 | 0.68 | 0.56 | 0.06 | 1.71 | 0.72 |
| SFA | 6.28 | 7.85 | 6.26 | 9.46 | 5.28 | 5.56 | 10.62 | 7.77 | 1.41 | 5.56 | 9.81 | 8.48 |
| PUFA | 3.09 | 3.93 | 3.53 | 4.87 | 2.99 | 2.81 | 5.02 | 3.73 | 0.62 | 2.84 | 4.87 | 3.89 |
| TFA | 0.36 | 0.29 | 0.46 | 0.31 | 0.34 | 0.28 | 0.29 | 0.28 | 0.07 | 0.14 | 0.38 | 0.27 |
| Fruits | 2.93 | 3.55 | 3.77 | 2.45 | 2.58 | 2.01 | 2.41 | 2.88 | 0.61 | 2.00 | 3.88 | 2.99 |
| Vegetables | 2.89 | 3.97 | 2.99 | 2.68 | 3.26 | 4.29 | 1.73 | 3.88 | 1.15 | 1.87 | 5.51 | 4.08 |
| Nuts and seeds | 0.32 | 0.39 | 0.38 | 0.32 | 0.26 | 0.27 | 0.33 | 0.33 | 0.14 | 0.13 | 0.59 | 0.37 |
| Whole grains | 0.35 | 0.74 | 0.06 | 0.01 | 0.11 | 0.36 | 0.27 | 0.39 | 0.33 | 0.01 | 0.85 | 0.45 |
| Legumes | 0.68 | 1.18 | 0.55 | 2.30 | 0.95 | 0.78 | 1.61 | 1.39 | 0.52 | 0.68 | 2.30 | 1.49 |
| Rice | 10.38 | 6.31 | 11.98 | 3.53 | 5.90 | 10.30 | 9.13 | 8.26 | 2.25 | 4.31 | 11.98 | 8.62 |
| Sweet potatoes | 0.12 | 0.11 | 0.03 | 0.18 | 0.12 | 0.10 | 0.18 | 0.13 | 0.08 | 0.03 | 0.29 | 0.10 |
| Physical activity | 8.54 | 8.15 | 7.04 | 8.47 | 12.49 | 7.95 | 5.06 | 7.48 | 3.10 | 2.63 | 12.49 | 7.45 |
| Child underweight | 19.16 | 19.70 | 22.65 | 22.31 | 28.86 | 24.16 | 12.46 | 18.18 | 6.77 | 5.47 | 27.27 | 20.38 |
| Sex M=1, F=2 | 10.56 | 6.31 | 5.80 | 7.13 | 6.59 | 6.62 | 10.47 | 9.21 | 5.20 | 4.29 | 19.02 | 6.02 |
| Total % weights | 79.92 | 80.01 | 74.68 | 81.34 | 81.42 | 80.63 | 79.83 | 80.74 | 2.17 | 76.96 | 81.91 | 80.96 |

**Table 3.** Consistency measured with 37 subgroups of worldwide GBD data.
4. Dose-response (Biological gradient) score=5 for the mean of absolute differences of BMI formula outputs and mean BMI for the dose-response quartiles< 0.300 kg/M^2^—Table 3 also shows that the mean of the BMI absolute differences between the BMI formula estimates and mean BMIs in the four dose-response quartiles averaged 0.220 kg/M^2^.
5. Temporality score=5 for the BMI trend formula versus BMI trend r>0.500, *p*<0.0001—Table 4 shows the mean trends the r values and the R^2^ values for each of the 22 risk factors. As is shown in Supplementary Table 6 and detailed in Appendix 1, a multiple regression analysis with the worldwide BMI trend 1990-2007 (dependent variable) versus 22 of the 23 risk factor mean trends and R^2^ values from Table 4 (independent variables excluding sex) generated a BMI trend formula (All foods are in Kcal/day.):

| Subsets of worldwide data on risk factors related to BMI | Final BMI formula n | Mean BMI | BMI formula output | Absolute differences | RCT 1-20 Mean | 80% CI low | 80% CI high | BMI output out of 80% CI |
| --- | --- | --- | --- | --- | --- | --- | --- | --- |
| SDI quartile 1 | 1926 | 24.486 | 24.496 | 0.010 | 24.455 | 24.138 | 24.738 |  |
| SDI quartile 2 | 2096 | 21.827 | 22.158 | 0.331 | 22.185 | 21.900 | 22.523 | 1 |
| SDI quartile 3 | 2220 | 20.401 | 19.931 | 0.471 | 19.976 | 19.584 | 20.338 | 1 |
| SDI quartile 4 | 1604 | 20.306 | 20.546 | 0.240 | 20.554 | 20.329 | 20.903 | 1 |
| Africa | 1682 | 21.666 | 21.429 | 0.237 | 21.447 | 21.121 | 21.772 |  |
| Asia | 4188 | 20.478 | 20.492 | 0.014 | 20.515 | 20.180 | 20.780 |  |
| Europe | 880 | 24.313 | 24.604 | 0.292 | 24.557 | 24.356 | 24.730 | 1 |
| North America | 558 | 25.912 | 25.494 | 0.418 | 25.477 | 25.016 | 25.810 | 1 |
| Oceania | 54 | 24.433 | 25.205 | 0.772 | 25.033 | 24.721 | 25.430 | 1 |
| South America | 468 | 23.463 | 24.128 | 0.665 | 24.222 | 23.788 | 24.594 | 1 |
| Four countries with subnational data | 730 | 24.848 | 24.795 | 0.052 | 24.775 | 24.296 | 25.110 |  |
| Quartile 1 BMI increasing foods | 1967 | 24.089 | 24.365 | 0.275 | 24.362 | 24.120 | 24.584 | 1 |
| Q2 | 1834 | 21.962 | 21.961 | 0.000 | 21.943 | 21.639 | 22.290 |  |
| Q3 | 2050 | 20.607 | 20.167 | 0.440 | 20.188 | 19.885 | 20.462 | 1 |
| Q4 | 1995 | 20.494 | 20.682 | 0.188 | 20.738 | 20.446 | 21.066 |  |
| Quartile 1 BMI decreasing foods | 1475 | 20.612 | 20.273 | 0.339 | 20.312 | 19.955 | 20.650 |  |
| Q2 | 2275 | 21.125 | 21.049 | 0.076 | 21.056 | 20.731 | 21.345 |  |
| Q3 | 2171 | 22.405 | 22.377 | 0.028 | 22.410 | 22.117 | 22.772 |  |
| Q4 | 1925 | 22.695 | 23.084 | 0.389 | 23.067 | 22.846 | 23.411 |  |
| Quartile 1 physical activity | 2371 | 20.480 | 20.311 | 0.169 | 20.314 | 20.062 | 20.555 |  |
| Q2 | 1573 | 21.663 | 21.881 | 0.217 | 21.912 | 21.658 | 22.230 |  |
| Q3 | 1998 | 21.325 | 21.324 | 0.001 | 21.343 | 21.026 | 21.727 |  |
| Q4 | 1904 | 23.923 | 23.963 | 0.039 | 23.974 | 23.553 | 24.369 |  |
| Male | 3923 | 21.485 | 21.420 | 0.065 | 21.388 | 21.138 | 21.566 |  |
| Female | 3923 | 22.051 | 22.120 | 0.068 | 22.180 | 21.858 | 22.501 |  |
| Bootstrap trial 1 | 100 | 21.715 | 21.741 | 0.025 | 21.755 | 21.503 | 22.013 |  |
| Bootstrap trial 2 | 100 | 22.040 | 21.986 | 0.055 | 22.011 | 21.724 | 22.254 |  |
| Bootstrap trial 3 | 100 | 21.676 | 21.744 | 0.068 | 21.769 | 21.479 | 22.019 |  |
| Bootstrap trial 4 | 100 | 21.926 | 21.944 | 0.018 | 21.954 | 21.659 | 22.206 |  |
| UK | 66 | 24.994 | 25.245 | 0.250 | 25.155 | 24.739 | 25.497 |  |
| USA | 376 | 26.017 | 25.809 | 0.208 | 25.684 | 25.177 | 26.352 |  |
| Japan | 158 | 21.892 | 22.888 | 0.996 | 22.898 | 22.254 | 23.327 | 1 |
| Mexico | 130 | 24.985 | 23.953 | 1.032 | 24.222 | 23.679 | 24.977 | 1 |
| Quartile 1 Dose-response: BMI formula versus BMI | 1928 | 24.763 | 24.949 | 0.186 | 24.936 | 24.617 | 25.230 |  |
| Q2 | 1469 | 22.568 | 22.480 | 0.088 | 22.525 | 22.187 | 22.805 |  |
| Q3 | 2647 | 20.872 | 21.069 | 0.197 | 21.069 | 20.884 | 21.325 | 1 |
| Q4 | 1802 | 19.227 | 18.818 | 0.409 | 18.859 | 18.443 | 19.220 | 1 |
| Mean of BMI - BMI formula absolute val |  |  |  | 0.252 |  |  |  |  |
| Mean of BMI-BMI formula dose response |  |  |  | 0.220 |  |  |  |  |
| Total out of 80% CI range |  |  |  |  |  |  |  | 13 |

**Table 4.** Trends of BMI and the BMI risk factors (1990-2017)

| **BMI trend versus risk factor trends, n=7846 cohorts** | **Mean** | **SD** | **Min** | **Max** | **R** | **95% CI** | **95% CI** | **P** | **R<sup>2</sup>** |
| --- | --- | --- | --- | --- | --- | --- | --- | --- | --- |
| **BMI trend** | 0.076 | 0.031 | -0.018 | 0.216 |  |  |  |  |  |
| **Processed meat Kcal/day trend** | -0.051 | 0.156 | -0.812 | 1.680 | -0.009 | -0.031 | 0.013 | 0.44 | 0.000 |
| **Red meat Kcal/day trend** | 0.556 | 1.084 | -3.291 | 4.731 | 0.410 | 0.392 | 0.428 | <.0001 | 0.168 |
| **Fish Kcal/day trend** | -0.224 | 0.953 | -0.130 | 9.790 | -0.158 | -0.180 | -0.137 | <.0001 | 0.025 |
| **Milk Kcal/day trend** | 0.190 | 0.299 | -2.018 | 2.503 | 0.206 | 0.185 | 0.227 | <.0001 | 0.043 |
| **Poultry Kcal/day trend** | 1.146 | 1.042 | -0.871 | 7.655 | 0.420 | 0.402 | 0.439 | <.0001 | 0.177 |
| **Eggs Kcal/day trend** | 0.300 | 0.492 | -0.877 | 1.275 | 0.484 | 0.467 | 0.501 | <.0001 | 0.234 |
| **Alcohol Kcal/day trend** | 0.031 | 0.164 | -0.914 | 1.993 | 0.289 | 0.268 | 0.309 | <.0001 | 0.083 |
| **Sugary beverages Kcal/day trend** | 0.289 | 1.139 | -5.401 | 14.001 | 0.125 | 0.103 | 0.147 | <.0001 | 0.016 |
| **Corn Kcal/day available trend** | -0.028 | 0.606 | -4.355 | 2.809 | 0.141 | 0.119 | 0.162 | <.0001 | 0.020 |
| **Potatoes Kcal/day available trend** | 0.246 | 1.837 | -12.59 | 11.031 | 0.272 | 0.251 | 0.292 | <.0001 | 0.074 |
| **SFA Kcal/day trend** | 0.210 | 0.925 | -3.660 | 2.095 | -0.050 | -0.072 | -0.028 | <.0001 | 0.002 |
| **PUFA Kcal/day trend** | 0.996 | 0.959 | -3.026 | 5.473 | 0.286 | 0.266 | 0.306 | <.0001 | 0.082 |
| **TFA kcal/day trend** | -0.210 | 0.446 | -2.681 | 0.013 | -0.066 | -0.088 | -0.044 | <.0001 | 0.004 |
| **Fruits Kcal/day trend** | 0.623 | 0.604 | -2.078 | 4.104 | 0.481 | 0.464 | 0.498 | <.0001 | 0.232 |
| **Vegetables Kcal/day trend** | 1.399 | 1.614 | -7.961 | 8.465 | 0.396 | 0.377 | 0.415 | <.0001 | 0.157 |
| **Nuts and seeds Kcal/day trend** | 0.276 | 0.265 | -0.338 | 3.124 | 0.123 | 0.101 | 0.144 | <.0001 | 0.015 |
| **Whole grains Kcal/day trend** | 0.173 | 0.586 | -2.604 | 8.463 | 0.104 | 0.082 | 0.126 | <.0001 | 0.011 |
| **Legumes Kcal/day trend** | 0.370 | 0.979 | -3.240 | 5.401 | -0.298 | -0.318 | -0.277 | <.0001 | 0.089 |
| **Rice Kcal/day available trend** | -0.787 | 1.292 | -5.051 | 3.496 | -0.393 | -0.412 | -0.374 | <.0001 | 0.155 |
| **Sweet potatoes Kcal/day available trend** | -0.460 | 1.339 | -4.818 | 3.905 | -0.499 | -0.516 | -0.482 | <.0001 | 0.249 |
| **Physical activity METs trend** | 4.724 | 6.826 | -33.86 | 28.629 | 0.379 | 0.360 | 0.398 | <.0001 | 0.144 |
| **Child underweight >2SD trend** | -0.004 | 0.004 | -0.014 | 0.001 | 0.045 | 0.023 | 0.067 | <.0001 | 0.002 |

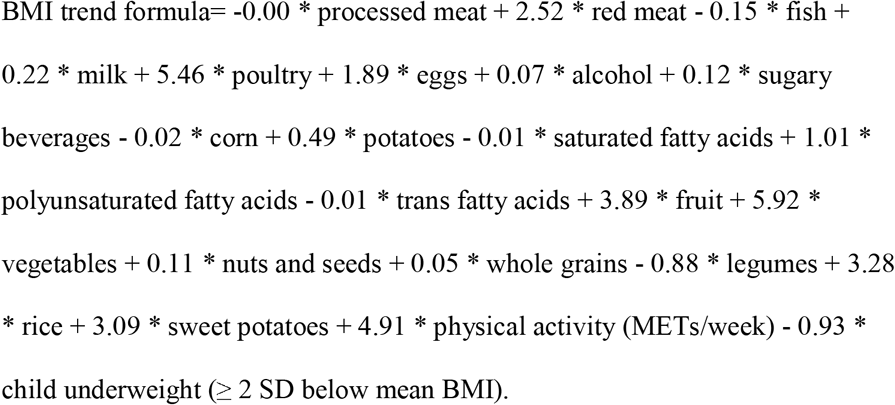 As with the BMI formula, the coefficients of the BMI risk factor trends formula were equated to their PAFs. BMI trend formula r=0.592 (H24 in Supplementary Table 6), 95% CI: 0.577 to 0.606, *p*<0.0001, and total PAF=35.02%.
6. Analogy score=4—The three metabolic risk factors correlated with BMI to test analogy were systolic blood pressure (SBP), low density lipoprotein cholesterol (LDL-C), and fasting plasma glucose (FPG).
  - BMI correlated with SBP: r=0.102, 95% CI: 0.080 to 0.124, *p*<0.0001.
  - BMI correlated with FPG: r=0.558, 95% CI: 0.542 to 0.573, *p*<0.0001.
  - BMI correlated with LDL-C: r=0.756, 95% CI: 0.746 to 0.765, *p*<0.0001. Since BMI versus SBP r<0.500, Bradford Hill causality score=4.
7. Plausibility: Score=5—Based on systematic medical literature reviews, physical activity inversely correlated with BMI,^17^ and BMI directly correlated with intakes of sugary beverages,^18^ alcohol,^19^ and animal foods.^20^ The relationship of adult BMI with early childhood severe underweight has not been reported worldwide. Since people in poor countries with high infant/childhood malnutrition have fewer resources to obtain animal foods, fatty acids, and alcohol and more need for physical exercise than in developed countries, it is highly plausible that childhood severe underweight negatively correlated with lower BMI in adulthood.
8. Specificity: Score=5 for the BMI formula being unique—the BMI formula was specific to worldwide BMI and would have been different from risk factor formulas modeling SBP, FPG, LDL-C, or any health outcome.
9. Coherence: Score=5—As evidenced by the near perfect score 39/40 on the first eight criteria, the BMI formula accurately modeled worldwide mean BMI—total causality criteria score: 44/45.

Table 5 shows BMI formula estimates for various patterns of diet and/or other BMI formula risk factors. For instance, increasing physical activity by adding a run for 1 hour/day at six miles/hour on average along with decreasing BMI increasing foods by isocalorically shifting 25% of BMI increasing foods (Kcal/day) to BMI decreasing foods was projected to reduce the mean cohort BMI from 26.66 kg/M^2^ to mean BMI=22.26 kg/M^2^.

**Table 5.** BMI formula predictions with different diet or other risk factor scenarios β.

| **Risk factor scenario** | **BMI**<br>**kg/M<sup>2</sup>** | **BMI**<br>**kg/M<sup>2</sup>**<br>**predicted** | **80 %**<br>**CI low** | **80 %**<br>**CI**<br>**high** |
| --- | --- | --- | --- | --- |
| **World (n=7846 cohorts mean 1990-2017)** | 21.77 | 21.77 | 21.33 | 22.13 |
| **World with all children severely underweight** |  | 16.90 | 15.79 | 18.05 |
| **World with no children severely underweight** |  | 22.88 | 22.35 | 23.38 |
| **USA** | 26.67 | 26.08 | 25.30 | 26.43 |
| **USA 25% kcal shifted to BMI decreasing foods<sup>†</sup>** |  | 23.41 | 22.31 | 23.93 |
| **USA physical activity mean plus 1 hour/day run** |  | 24.94 | 24.05 | 25.67 |
| **USA physical activity mean + 1 hour run and 25% shift to BMI decreasing foods<sup>†</sup>** |  | 22.26 | 21.63 | 23.00 |
| **USA no red or processed meat** |  | 25.36 | 24.67 | 25.72 |
| **USA no sugary beverages** |  | 26.03 | 25.17 | 26.34 |
| **USA vegetarian** |  | 24.13 | 23.44 | 24.47 |
| **USA vegan** |  | 23.47 | 22.67 | 23.97 |
| **EAT-Lancet diet** |  | 22.86 | 21.67 | 23.53 |
| **Low Carb Mediterranean Diet** |  | 29.05 | 27.92 | 29.54 |

$\beta$ BMI formula estimates based on 28 years of following dietary and risk factor patterns
<sup>†</sup> Kcal/day 13 BMI increasing foods isocalorically shifted to the 7 BMI decreasing foods in the
BMI formula, distributed proportionally.

## Discussion

This paper’s analysis dataset formatted from GBD worldwide data used with the BMI formula enabled quantifying 23 risk factor PAFs for worldwide mean BMI for cohorts of 15-69-year-old people. This quantitatively rigorous methodology satisfied all nine Bradford Hill criteria for showing the BMI formula derived accurately modeled BMI.

The BMI formula PAFs are quite consistent with a 20-year-long observational study with 120,877 US male and female adults reported by Mozaffarian, et. al. They measured weight change per four-year period per 1.00 increased serving per day of various foods. It found the following foods associated with increased weights over four years: potato chips (0.77 kg), potatoes (0.58 kg), sugar-sweetened beverages (0.45 kg), unprocessed red meats (0.43 kg), and processed meats (0.42 kg). With the same methodology, the following foods associated with decreased weights: vegetables (−0.10 kg), whole grains (−0.17 kg), fruits (−0.22 kg), nuts (−0.26 kg), and yogurt (−0.37 kg) (*p*≤0.005 for each comparison).^21^

According to the International Potato Center, potato availability (a covariate), which positively correlated with BMI, included ≥50% highly processed potato products worldwide.^22^ This likely partially accounted for the positive correlation of potatoes with BMI. Higher intakes of ultra-processed foods overall have been associated with overweight/obesity.^23^ Ultra-processed food intakes have increased dramatically in developed countries in recent decades and are now increasing in middle-income countries.^24^

With the results of this paper consistent with and adding to previous findings, we should consider including the quantification of BMI risk factors demonstrated here into US Department of Health and Human Services (USDHHS)/ US Department of Agriculture (USDA) health policy discussions and clinical nutrition guideline deliberations. For instance, the Supplemental Nutrition Assistance Program (SNAP—formerly Food Stamps) spent an estimated 22.6% of its $73 billion/year budget in 2019^25^ on payments to low-income Americans for “sweetened beverages, prepared desserts, salty snacks, candy, and sugar.”^26^ Additionally, the USDA has subsidised crops that go primarily for animal feed or that are processed into sugars while not subsidising fruits and vegetables.^27^ While the USDA recognises the relatively low intake of fruits and vegetables in the USA and sponsors a publicity campaign to increase fruits and vegetables consumption,^28^ USDA expenditures should promote reduced prices of BMI decreasing foods and increased prices of BMI increasing foods.

A review of the literature on food costs relative to nutrient quality found that the median costs of fruits and vegetables (€0.82/100 kcal) were quite high relative meat/eggs/fish (€0.64/100 kcal), fresh dairy (€0.32/100 kcal), nuts (€0.25/100 kcal), and starches (€0.14/100 kcal).^29^ The economics of food, including the high cost of fruits and vegetables relative to animal foods, starchy potatoes, and sugary beverages, may contribute to overweight and obesity in the USA and in other affluent countries.

Following a low-carbohydrate, high-fat diet has been demonstrated to cause modest short term weight loss in obese people,^30^ However, the BMI formula projects that the long-term effect of a low-carb, high fat diet would be a mean cohort BMI of 29.05 kg/M^2^ 80% CI: 27.92 to 29.54 kg/M^2^ (Table 5).

### Limitations

The GBD data on animal foods, plant foods, alcohol, and fatty acids were not comprehensive and comprised only 1191.4 Kcal/day on average worldwide. Subnational data were available on only four countries. Because the data formatting and statistical methodology were new, this was necessarily a post hoc analysis and no pre-analysis protocol or Bradford Hill causality criteria scoring system was possible. As detailed in the Foresight Report on obesity,^31^ obesity is affected by a complex system of interacting factors besides diet, physical activity, and childhood feeding patterns, and sex. So genes,^32^ gut microbiome,^33^ ultra-processing of food,^34, 35^ and other influences on BMI were outside of the purview of this analysis.

### Generalisability

Over 70% of the cohorts in analysis dataset had low to moderate BMI levels (BMI < 23 kg/M^2^), so the results could be further refined for relatively high SDI countries. For results more applicable to high BMI countries, BMI formulas might also be derived from (1) the four countries that have subnational data (UK, USA, Mexico, and Japan n=730 cohorts, mean BMI=24.85 kg/M^2^) (2) the cohorts with mean BMI ≥ 23 kg/M^2^ (n=2256 cohorts, mean BMI=24.84 kg/M^2^), or (3) high SDI countries (n=1926 cohorts, mean BMI=24.49 kg/M^2^).

## Conclusions

Nine Bradford Hill causality criteria strongly supported that the BMI formula derived with 23 risk factors in proportion to their PAFs accurately modeled worldwide cohort mean BMI. While this study dealt only with dietary and other available risk factors for BMI (overweight/obesity), the AI methodology introduced could easily apply to estimating PAFs of multiple dietary and other risk factors that pertain to dozens of non-communicable disease health outcomes, for which the IHME have GBD data.

**Supplementary Table 1.** Definitions of IHME GBD risk factors and covariates related to BMI.

| Variables | Definition |
| --- | --- |
| **Alcohol** | Any alcohol consumption (g/day) |
| **Body-mass index** | Body mass index (BMI) ($\text{kg}/\text{m}^2$)—the dependent variable of interest |
| **Child underweight** | Proportion of children – 3 SD to – 2 SD of the WHO 2006 standard weight-for-age curve (0-1) |
| **Corn** | Corn availability percapita (g/day), a covariate |
| **Discontinued breast feeding** | Proportion of children aged 6-23 months who do not receive any breast milk |
| **Eggs** | Eggs availability percapita (g/day) a covariate |
| **Fasting plasma glucose** | Fasting plasma glucose (mmol/L) |
| **Fish** | This variable expressed in g/day was derived by determining the weight of fish in g corresponding to 1 g of omega-3 fatty acids (eicosapentaenoic acid and docosahexaenoic acid) by averaging the fish g per 1 g of omega-3 fatty acids 20 species of fish= 117.04 g/day fish/1 g/day omega-3 fatty acids (Supplementary Table 2) |
| **Fruits** | Consumption of fruits (includes fresh, frozen, cooked, canned, or dried fruit but excludes fruit juices and salted or pickled fruits) (g/day) |
| **Kilocalories available /day** | The mean number of kilocalories percapita available per day to people in each location (kcal/day available), a covariate |
| **LDL cholesterol** | Serum low-density lipoprotein cholesterol (mmol/L) |
| **Legumes** | Consumption of beans, lentils, pulses (g/day) |
| **Milk** | Consumption of milk including non-fat, low-fat, and full-fat milk but excluding soy milk and other plant derivatives (g/day) |
| **Nuts and seeds** | Consumption of nuts and seeds (g/day) |
| **Physical activity** | Average weekly physical activity at work, home, transport-related and recreational measured by MET min per week. Less than 3000 METs per week constitutes low physical activity. |
| **Poultry** | Poultry availability percapita (g/day), a covariate |
| **Potatoes** | Potatoes availability percapita (g/day), a covariate |
| **Processed meat** | Consumption of any processed meat (includes meat preserved by smoking, curing, salting, or addition of chemical preservatives, including bacon, salami, sausages, or deli or luncheon meats like ham, turkey, and pastrami (g/day) |
| **Red meat** | Consumption of red meat (includes beef, pork, lamb, and goat but excludes poultry, fish, eggs, and all processed meats) (g/day) |
| **Rice** | Rice availability percapita (g/day), a covariate |
| **Seafood omega-3 fatty acids** | Seafood omega-3 fatty acids (eicosapentaenoic acid and docosahexaenoic acid) in tablet or fish form (g/day) |
| **Socio-demographic index** | SDI is a composite indicator of development status that was originally constructed for GBD 2015, and is derived from components that correlate strongly with health outcomes. It is the geometric mean for indices of the total fertility rate among women younger than 25 years, mean education for those aged 15 years or older, and lag-distributed income per capita. The resulting metric ranges from 0 to 1, with higher values corresponding to higher levels of development. |
| **Sugar-sweetened beverages** | Consumption of any beverage with $\geq 50$ calories of sugar per one-cup serving, including carbonated beverages, sodas, energy drinks, fruit drinks but excluding 100% fruit and vegetable juices (g/day) |
| **Sweet potatoes** | Sweet potato availability percapita (g/day), a covariate |
| **Systolic blood pressure** | Systolic blood pressure (mm Hg) |
| **Total sugar** | Total sugar availability percapita (g/day), a covariate |
| **Vegetables** | Consumption of frozen, cooked, canned, or dried vegetables (including legumes but excluding salted or pickled, juices, nuts and seeds, and starchy vegetables such as potatoes or corn) (g/day) |
| **Whole grains** | Consumption of whole grains (bran, germ, and endosperm in their natural proportions) from breakfast cereals, bread, rice, pasta, biscuits, muffins, tortillas, pancakes, and others (g/day) |

**Supplementary Table 2.** Omega-3 Fatty Acid g to fish g calculation¶.

| **Fish** | **DHA**<br>**g/3**<br>**ounce**<br>**fish** | **EPA**<br>**g/3**<br>**ounce**<br>**fish** | **Omega-3 Fatty**<br>**Acids (DHA _**<br>**EPA) g/3 ounce**<br>**fish mean** | **Fish 3**<br>**ounces**<br>**= 85.02**<br>**g** | **Fish (g) per**<br>**omega-3**<br>**Fatty Acids**<br>**(g)=columns**<br>**E/F** |
| --- | --- | --- | --- | --- | --- |
| **Salmon Atlantic farmed** | 1.24 | 0.59 |  |  |  |
| **Salmon Atlantic wild** | 1.22 | 0.35 |  |  |  |
| **Herring Atlantic** | 0.94 | 0.77 |  |  |  |
| **Sardines canned in tomato**<br>**sauce drained** | 0.74 | 0.45 |  |  |  |
| **Mackerel Atlantic** | 0.59 | 0.43 |  |  |  |
| **Salmon pink canned**<br>**drained** | 0.63 | 0.28 |  |  |  |
| **Trout rainbow wild** | 0.44 | 0.40 |  |  |  |
| **Oysters eastern wild** | 0.23 | 0.30 |  |  |  |
| **Sea bass** | 0.47 | 0.18 |  |  |  |
| **Shrimp** | 0.12 | 0.12 |  |  |  |
| **Lobster** | 0.07 | 0.10 |  |  |  |
| **Tuna light canned in water**<br>**drained** | 0.17 | 0.02 |  |  |  |
| **Tilapia** | 0.11 |  |  |  |  |
| **Scallops** | 0.09 | 0.06 |  |  |  |
| **Cod Pacific** | 0.1 | 0.04 |  |  |  |
| **Tuna yellowfin** | 0.09 | 0.01 |  |  |  |
| **Mean DHA and EPA**<br>**Omega-3 Fatty Acids g/3**<br>**ounce fish** | 0.4531 | 0.2733 |  |  |  |
| **Calculations total Omega-3**<br>**FA g to fish g** |  |  | 0.7264 | 85.02 | 117.043 |

¶ Data on omega-3 fatty acid content of varieties of fish came from the National Institutes of Health Office of Dietary Supplements (USA)

**Supplementary Table 3.** Calculations of kcal/day from g/day of animal and plant foods¶.

| **Foods** | **Food sub-categories** | **kcal/serving** | **g/serving** | **kcal/g** |
| --- | --- | --- | --- | --- |
| **Milk (2% fat)** |  | 122 | 244 | 0.5 |
| **Fish** |  | 218 | 170 | 1.28 |
| **Eggs** |  | 72 | 50 | 1.44 |
| **Poultry** |  | 187 | 85 | 2.91 |
| **Red meat** |  | 247 | 85 | 2.91 |
| **Processed meat** |  |  |  |  |
|  | **Salami** | 222 | 59 | 3.76 |
|  | **Pastrami** | 104 | 71 | 1.46 |
|  | **Ring baloney** | 86 | 28 | 3.07 |
|  | **Pepperoni** | 94 | 100 | 0.94 |
| **Average processed meat** |  | 126.5 | 64.5 | 1.96 |
| **Fruits** |  | 97 | 162 | 0.60 |
| **Vegetables** |  | 59 | 91 | 0.65 |
| **Legumes** |  | 249 | 179 | 1.39 |
| **Nuts** |  | 172 | 28 | 6.14 |
| **Seeds** |  |  |  |  |
|  | **Flax seeds** | 55 | 10 | 5.5 |
|  | **Chia seeds** | 58 | 12 | 4.83 |
|  | **Fennel seeds** | 34.5 | 10 | 3.45 |
|  | **Hemp seeds** | 55.3 | 10 | 5.53 |
| **Average of seeds** |  | 50.7 | 10.5 | 4.83 |
| **Average of nuts and seeds** |  | 111.4 | 19.25 | 5.78 |
| **Corn** |  | 99 | 103 | 0.96 |
| **Potatoes** |  | 161 | 173 | 0.93 |
| **Sweet potatoes** |  | 115 | 151 | 0.76 |
| **Rice** |  | 205 | 158 | 1.3 |
| **Whole grains** |  | 120 | 52 | 2.31 |

¶ Source: NutritionIX app

**Supplementary Table 4.** Pearson correlations between BMI formula risk factors from Table 1.

| Correlations between BMI risk factors (r) | Red meat | Fish | Milk | Poultry | Eggs | Alcohol | Sugary beverages | Corn | Potatoes | SFA | PUFA |
| --- | --- | --- | --- | --- | --- | --- | --- | --- | --- | --- | --- |
| Processed meat KC/d | 0.647 | 0.286 | 0.756 | 0.662 | 0.569 | 0.576 | -0.108 | -0.050 | 0.159 | 0.624 | 0.699 |
| Red meat KC/d | 1.000 | 0.118 | 0.676 | 0.664 | 0.784 | 0.466 | 0.046 | -0.143 | 0.123 | 0.704 | 0.694 |
| Fish KC/d | 0.118 | 1.000 | 0.109 | 0.163 | 0.393 | 0.294 | -0.163 | -0.018 | -0.058 | 0.038 | 0.163 |
| Milk KC/d | 0.676 | 0.109 | 1.000 | 0.673 | 0.525 | 0.385 | 0.072 | -0.107 | 0.202 | 0.766 | 0.680 |
| Poultry KC/d | 0.664 | 0.163 | 0.673 | 1.000 | 0.628 | 0.351 | 0.142 | 0.011 | 0.102 | 0.701 | 0.837 |
| Eggs KC/d | 0.784 | 0.393 | 0.525 | 0.628 | 1.000 | 0.341 | 0.021 | -0.099 | 0.030 | 0.595 | 0.631 |
| Alcohol KC/d | 0.466 | 0.294 | 0.385 | 0.351 | 0.341 | 1.000 | -0.090 | -0.107 | -0.011 | 0.287 | 0.379 |
| Sugary beverages KC/d | 0.046 | -0.163 | 0.072 | 0.142 | 0.021 | -0.090 | 1.000 | 0.419 | -0.025 | 0.045 | 0.113 |
| Corn KC/d | -0.143 | -0.018 | -0.107 | 0.011 | -0.099 | -0.107 | 0.419 | 1.000 | 0.158 | -0.133 | -0.034 |
| Potatoes KC/d | 0.123 | -0.058 | 0.202 | 0.102 | 0.030 | -0.011 | -0.025 | 0.158 | 1.000 | 0.211 | 0.086 |
| SFA KC/d | 0.704 | 0.038 | 0.766 | 0.701 | 0.595 | 0.287 | 0.045 | -0.133 | 0.211 | 1.000 | 0.635 |
| PUFAs KC/d | 0.694 | 0.163 | 0.680 | 0.837 | 0.631 | 0.379 | 0.113 | -0.034 | 0.086 | 0.635 | 1.000 |
| TFA KC/d | 0.187 | -0.008 | 0.428 | 0.446 | 0.203 | 0.152 | 0.198 | 0.269 | -0.091 | 0.233 | 0.433 |
| Fruits KC/d | 0.371 | 0.098 | 0.490 | 0.606 | 0.366 | 0.058 | 0.198 | 0.102 | 0.243 | 0.470 | 0.589 |
| Vegetables KC/d | 0.382 | 0.272 | 0.349 | 0.345 | 0.513 | 0.191 | -0.167 | -0.203 | -0.047 | 0.224 | 0.385 |
| Nuts and seeds KC/d | 0.386 | 0.072 | 0.579 | 0.504 | 0.333 | 0.232 | -0.146 | -0.112 | 0.187 | 0.471 | 0.527 |
| Whole grains KC/d | -0.112 | 0.091 | -0.254 | -0.023 | -0.077 | 0.066 | 0.204 | 0.428 | -0.058 | -0.165 | -0.063 |
| Legumes KC/d | -0.404 | 0.061 | -0.194 | -0.220 | -0.416 | 0.140 | 0.197 | 0.269 | 0.019 | -0.389 | -0.145 |
| Rice KC/d | -0.351 | -0.070 | -0.565 | -0.389 | -0.271 | -0.129 | -0.148 | -0.250 | -0.377 | -0.378 | -0.368 |
| Sweet potatoes KC/d | 0.005 | -0.074 | -0.395 | -0.240 | 0.049 | -0.063 | -0.218 | -0.028 | 0.239 | -0.130 | -0.234 |
| Physical activity METs | -0.068 | -0.163 | -0.430 | -0.396 | -0.114 | 0.105 | -0.103 | 0.028 | 0.022 | -0.309 | -0.431 |
| Child underweight | -0.736 | -0.189 | -0.403 | -0.618 | -0.803 | -0.131 | -0.065 | -0.083 | -0.215 | -0.584 | -0.622 |
| Correlations between BMI risk factors (r) | TFA | Fruits | Vegetables | Nuts-seeds | Whole grains | Legumes | Rice | Sweet potatoes | Physical activity | Child underweight | Sex m/f |
| Processed meat KC/d | 0.463 | 0.379 | 0.290 | 0.504 | -0.023 | -0.197 | -0.391 | -0.194 | -0.212 | -0.411 | -0.086 |
| Red meat KC/d | 0.187 | 0.371 | 0.382 | 0.386 | -0.112 | -0.404 | -0.351 | 0.005 | -0.068 | -0.736 | -0.230 |
| Fish KC/d | -0.008 | 0.098 | 0.272 | 0.072 | 0.091 | 0.061 | -0.070 | -0.074 | -0.163 | -0.189 | -0.023 |
| Milk KC/d | 0.428 | 0.490 | 0.349 | 0.579 | -0.254 | -0.194 | -0.565 | -0.395 | -0.430 | -0.403 | -0.018 |
| Poultry KC/d | 0.446 | 0.606 | 0.345 | 0.504 | -0.023 | -0.220 | -0.389 | -0.240 | -0.396 | -0.618 | 0.000 |
| Eggs KC/d | 0.203 | 0.366 | 0.513 | 0.333 | -0.077 | -0.416 | -0.271 | 0.049 | -0.114 | -0.803 | 0.000 |
| Alcohol KC/d | 0.152 | 0.058 | 0.191 | 0.232 | 0.066 | 0.140 | -0.129 | -0.063 | 0.105 | -0.131 | -0.432 |
| Sugary beverages KC/d | 0.198 | 0.198 | -0.167 | -0.146 | 0.204 | 0.197 | -0.148 | -0.218 | -0.103 | -0.065 | -0.311 |
| Corn KC/d | 0.269 | 0.102 | -0.203 | -0.112 | 0.428 | 0.269 | -0.250 | -0.028 | 0.028 | -0.083 | 0.000 |
| Potatoes KC/d | -0.091 | 0.243 | -0.047 | 0.187 | -0.058 | 0.019 | -0.377 | 0.239 | 0.022 | -0.215 | 0.000 |
| SFA KC/d | 0.233 | 0.470 | 0.224 | 0.471 | -0.165 | -0.389 | -0.378 | -0.130 | -0.309 | -0.584 | 0.007 |
| PUFAs KC/d | 0.433 | 0.589 | 0.385 | 0.527 | -0.063 | -0.145 | -0.368 | -0.234 | -0.431 | -0.622 | -0.013 |
| TFA KC/d | 1.000 | 0.387 | 0.284 | 0.312 | 0.087 | 0.044 | -0.261 | -0.322 | -0.412 | -0.121 | 0.096 |
| Fruits KC/d | 0.387 | 1.000 | 0.386 | 0.579 | 0.012 | -0.028 | -0.319 | -0.096 | -0.391 | -0.493 | 0.116 |
| Vegetables KC/d | 0.284 | 0.386 | 1.000 | 0.547 | -0.349 | -0.157 | -0.230 | -0.095 | -0.208 | -0.453 | -0.066 |
| Nuts and seeds KC/d | 0.312 | 0.579 | 0.547 | 1.000 | -0.211 | -0.023 | -0.352 | -0.074 | -0.302 | -0.293 | -0.030 |
| Whole grains KC/d | 0.087 | 0.012 | -0.349 | -0.211 | 1.000 | 0.100 | 0.548 | -0.039 | 0.268 | 0.089 | -0.064 |
| Legumes KC/d | 0.044 | -0.028 | -0.157 | -0.023 | 0.100 | 1.000 | -0.050 | -0.054 | -0.068 | 0.506 | -0.142 |
| Rice KC/d | -0.261 | -0.319 | -0.230 | -0.352 | 0.548 | -0.050 | 1.000 | 0.070 | 0.386 | 0.374 | 0.000 |
| Sweet potatoes KC/d | -0.322 | -0.096 | -0.095 | -0.074 | -0.039 | -0.054 | 0.070 | 1.000 | 0.327 | -0.167 | 0.000 |
| Physical activity | -0.412 | -0.391 | -0.208 | -0.302 | 0.268 | -0.068 | 0.386 | 0.327 | 1.000 | 0.135 | -0.257 |
| Child underweight | -0.121 | -0.493 | -0.453 | -0.293 | 0.089 | 0.506 | 0.374 | -0.167 | 0.135 | 1.000 | -0.017 |

**Red: Fruits, vegetables, and nuts and seeds correlated positively with BMI increasing foods**
**Blue: Fruits, vegetables, and nuts and seeds correlated positively with themselves**
**Green: Fruits, vegetables, and nuts and seeds correlated negatively with BMI decreasing foods**

**Supplementary Table 5.**
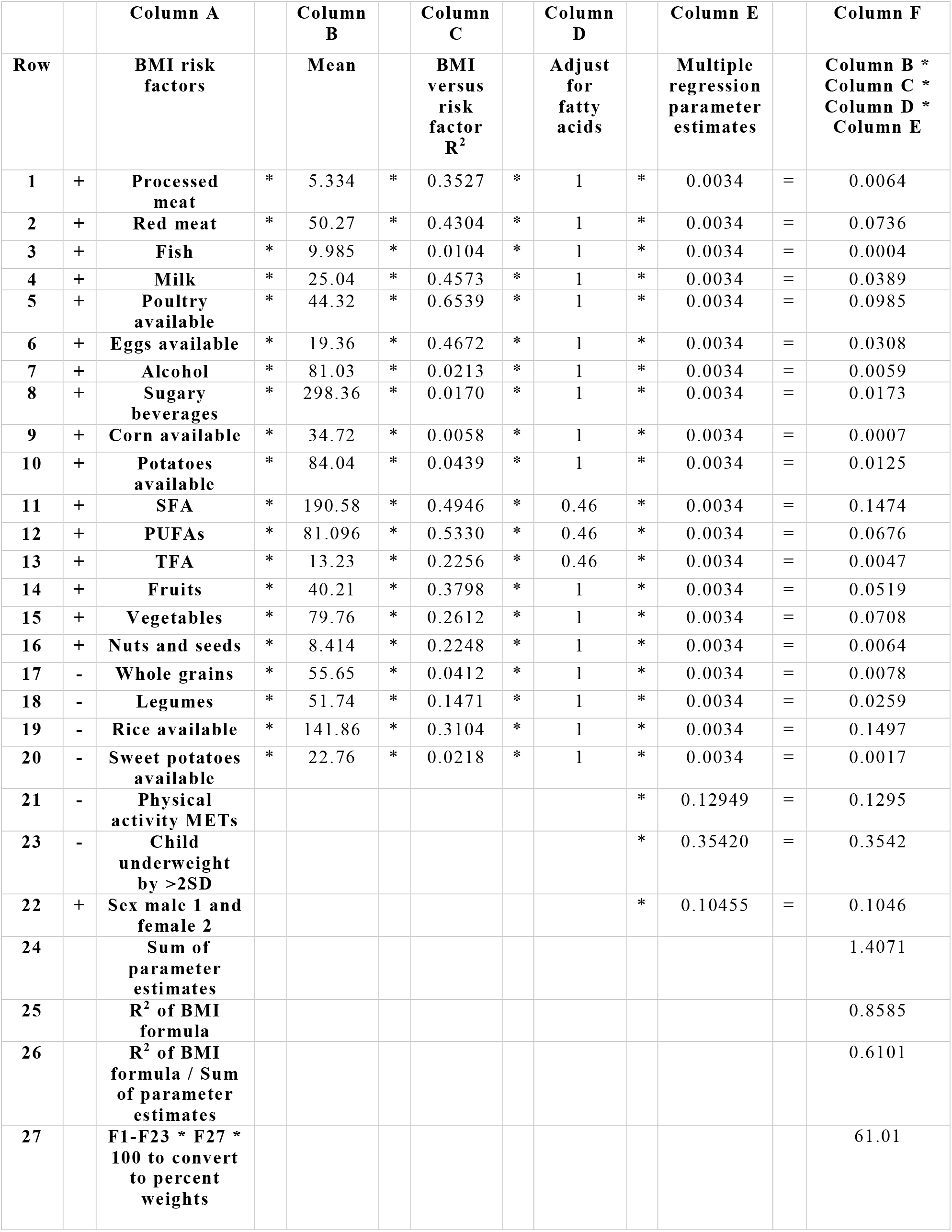
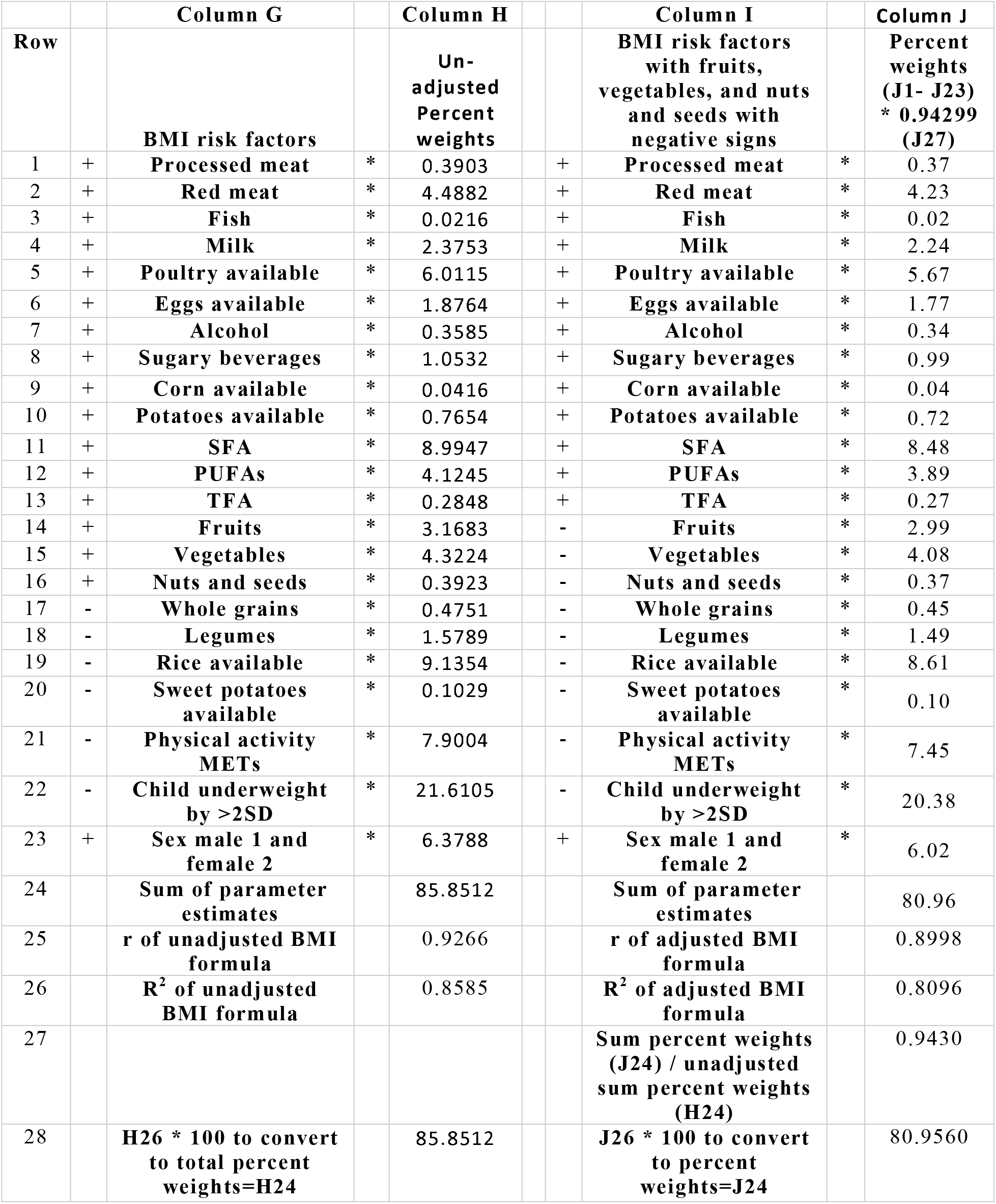
Derivation of the BMI formula.

|  |  | Column A |  | Column B |  | Column C |  | Column D |  | Column E |  | Column F |
| --- | --- | --- | --- | --- | --- | --- | --- | --- | --- | --- | --- | --- |
| Row | | BMI risk factors | | Mean | | BMI versus risk factor $R^2$ | | Adjust for fatty acids | | Multiple regression parameter estimates | | Column B *<br>Column C *<br>Column D *<br>Column E |
| 1 | + | Processed meat | * | 5.334 | * | 0.3527 | * | 1 | * | 0.0034 | = | 0.0064 |
| 2 | + | Red meat | * | 50.27 | * | 0.4304 | * | 1 | * | 0.0034 | = | 0.0736 |
| 3 | + | Fish | * | 9.985 | * | 0.0104 | * | 1 | * | 0.0034 | = | 0.0004 |
| 4 | + | Milk | * | 25.04 | * | 0.4573 | * | 1 | * | 0.0034 | = | 0.0389 |
| 5 | + | Poultry available | * | 44.32 | * | 0.6539 | * | 1 | * | 0.0034 | = | 0.0985 |
| 6 | + | Eggs available | * | 19.36 | * | 0.4672 | * | 1 | * | 0.0034 | = | 0.0308 |
| 7 | + | Alcohol | * | 81.03 | * | 0.0213 | * | 1 | * | 0.0034 | = | 0.0059 |
| 8 | + | Sugary beverages | * | 298.36 | * | 0.0170 | * | 1 | * | 0.0034 | = | 0.0173 |
| 9 | + | Corn available | * | 34.72 | * | 0.0058 | * | 1 | * | 0.0034 | = | 0.0007 |
| 10 | + | Potatoes available | * | 84.04 | * | 0.0439 | * | 1 | * | 0.0034 | = | 0.0125 |
| 11 | + | SFA | * | 190.58 | * | 0.4946 | * | 0.46 | * | 0.0034 | = | 0.1474 |
| 12 | + | PUFAs | * | 81.096 | * | 0.5330 | * | 0.46 | * | 0.0034 | = | 0.0676 |
| 13 | + | TFA | * | 13.23 | * | 0.2256 | * | 0.46 | * | 0.0034 | = | 0.0047 |
| 14 | + | Fruits | * | 40.21 | * | 0.3798 | * | 1 | * | 0.0034 | = | 0.0519 |
| 15 | + | Vegetables | * | 79.76 | * | 0.2612 | * | 1 | * | 0.0034 | = | 0.0708 |
| 16 | + | Nuts and seeds | * | 8.414 | * | 0.2248 | * | 1 | * | 0.0034 | = | 0.0064 |
| 17 | - | Whole grains | * | 55.65 | * | 0.0412 | * | 1 | * | 0.0034 | = | 0.0078 |
| 18 | - | Legumes | * | 51.74 | * | 0.1471 | * | 1 | * | 0.0034 | = | 0.0259 |
| 19 | - | Rice available | * | 141.86 | * | 0.3104 | * | 1 | * | 0.0034 | = | 0.1497 |
| 20 | - | Sweet potatoes available | * | 22.76 | * | 0.0218 | * | 1 | * | 0.0034 | = | 0.0017 |
| 21 | - | Physical activity METs |  |  |  |  |  |  | * | 0.12949 | = | 0.1295 |
| 23 | - | Child underweight by >2SD |  |  |  |  |  |  | * | 0.35420 | = | 0.3542 |
| 22 | + | Sex male 1 and female 2 |  |  |  |  |  |  | * | 0.10455 | = | 0.1046 |
| 24 |  | Sum of parameter estimates |  |  |  |  |  |  |  |  |  | 1.4071 |
| 25 | | $R^2$ of BMI formula | | | | | | | | | | 0.8585 |
| 26 | | $R^2$ of BMI formula / Sum of parameter estimates | | | | | | | | | | 0.6101 |
| 27 |  | F1-F23 * F27 * 100 to convert to percent weights |  |  |  |  |  |  |  |  |  | 61.01 |

**Supplementary Table 5. Derivation of the BMI formula continued**
| Row |  | Column G |  | Column H |  | Column I |  | Column J |
| --- | --- | --- | --- | --- | --- | --- | --- | --- |
|  |  | BMI risk factors |  | Un-adjusted Percent weights |  | BMI risk factors with fruits, vegetables, and nuts and seeds with negative signs |  | Percent weights (J1- J23) * 0.94299 (J27) |
| 1 | + | Processed meat | * | 0.3903 | + | Processed meat | * | 0.37 |
| 2 | + | Red meat | * | 4.4882 | + | Red meat | * | 4.23 |
| 3 | + | Fish | * | 0.0216 | + | Fish | * | 0.02 |
| 4 | + | Milk | * | 2.3753 | + | Milk | * | 2.24 |
| 5 | + | Poultry available | * | 6.0115 | + | Poultry available | * | 5.67 |
| 6 | + | Eggs available | * | 1.8764 | + | Eggs available | * | 1.77 |
| 7 | + | Alcohol | * | 0.3585 | + | Alcohol | * | 0.34 |
| 8 | + | Sugary beverages | * | 1.0532 | + | Sugary beverages | * | 0.99 |
| 9 | + | Corn available | * | 0.0416 | + | Corn available | * | 0.04 |
| 10 | + | Potatoes available | * | 0.7654 | + | Potatoes available | * | 0.72 |
| 11 | + | SFA | * | 8.9947 | + | SFA | * | 8.48 |
| 12 | + | PUFAs | * | 4.1245 | + | PUFAs | * | 3.89 |
| 13 | + | TFA | * | 0.2848 | + | TFA | * | 0.27 |
| 14 | + | Fruits | * | 3.1683 | - | Fruits | * | 2.99 |
| 15 | + | Vegetables | * | 4.3224 | - | Vegetables | * | 4.08 |
| 16 | + | Nuts and seeds | * | 0.3923 | - | Nuts and seeds | * | 0.37 |
| 17 | - | Whole grains | * | 0.4751 | - | Whole grains | * | 0.45 |
| 18 | - | Legumes | * | 1.5789 | - | Legumes | * | 1.49 |
| 19 | - | Rice available | * | 9.1354 | - | Rice available | * | 8.61 |
| 20 | - | Sweet potatoes available | * | 0.1029 | - | Sweet potatoes available | * | 0.10 |
| 21 | - | Physical activity METs | * | 7.9004 | - | Physical activity METs | * | 7.45 |
| 22 | - | Child underweight by >2SD | * | 21.6105 | - | Child underweight by >2SD | * | 20.38 |
| 23 | + | Sex male 1 and female 2 | * | 6.3788 | + | Sex male 1 and female 2 | * | 6.02 |
| 24 |  | Sum of parameter estimates |  | 85.8512 |  | Sum of parameter estimates |  | 80.96 |
| 25 |  | r of unadjusted BMI formula |  | 0.9266 |  | r of adjusted BMI formula |  | 0.8998 |
| 26 |  | R<sup>2</sup> of unadjusted BMI formula |  | 0.8585 |  | R<sup>2</sup> of adjusted BMI formula |  | 0.8096 |
| 27 |  |  |  |  |  | Sum percent weights (J24) / unadjusted sum percent weights (H24) |  | 0.9430 |
| 28 |  | H26 * 100 to convert to total percent weights=H24 |  | 85.8512 |  | J26 * 100 to convert to percent weights=J24 |  | 80.9560 |

**Supplementary Table 6.**
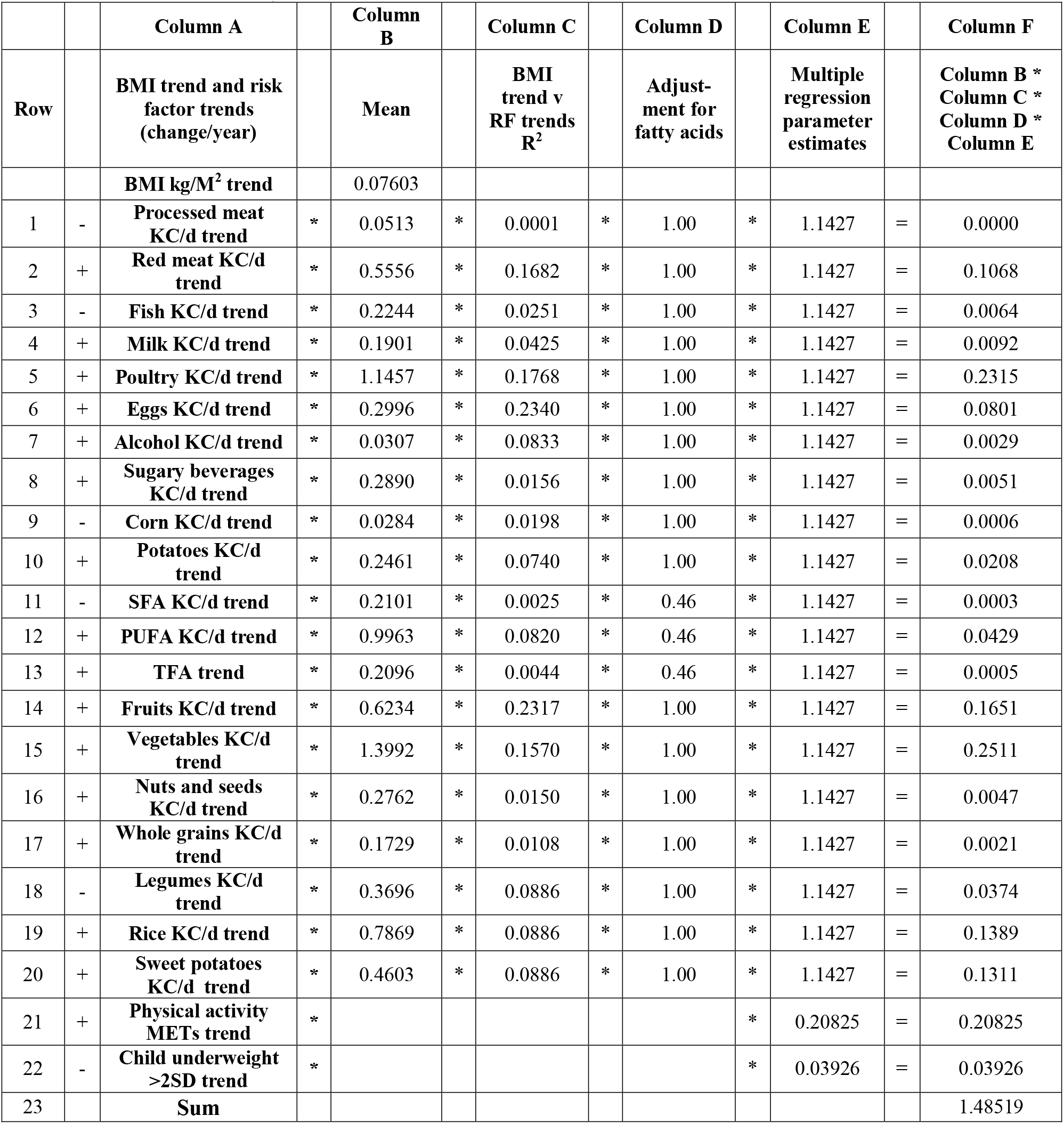
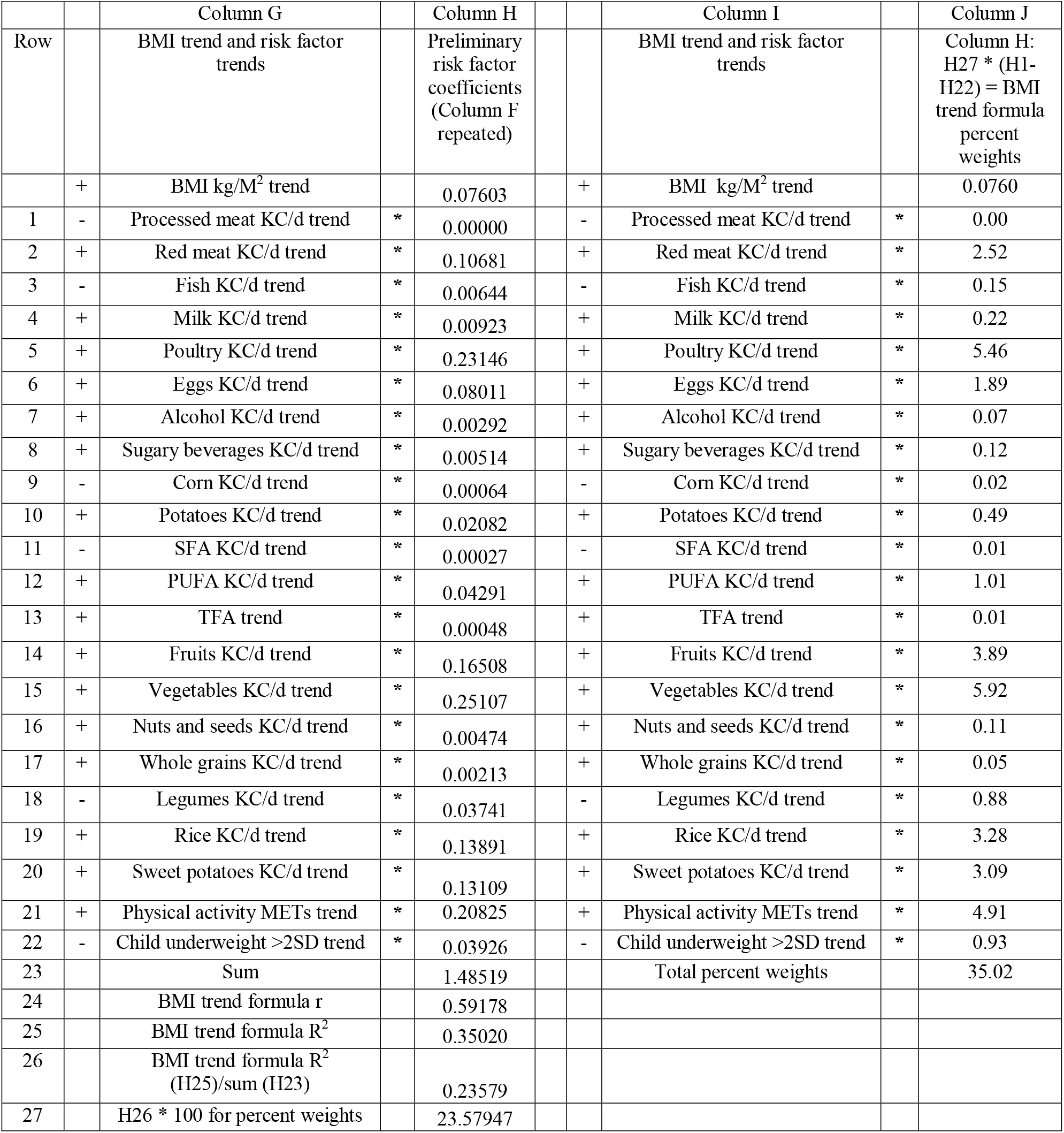
BMI trend versus risk factor trends formula derivation (all variables standardized)

## Data Availability

The raw, unformatted data used in this analysis is now out of date. The 2019 GBD data on all the variables in this analysis may be obtained from the IHME by volunteer collaborating researchers. The formatted database, SAS codes and Excel spreadsheets on which this analysis is based are posted on the Mendeley data repository.

https://data.mendeley.com/v1/datasets/publish-confirmation/g6b39zxck4/6

## Article Information

### Corresponding Author

David K. Cundiff, Independent researcher

### contributions

DKC acts as guarantor; conceived and designed the study, acquired and analysed the data, interpreted the study findings, drafted the manuscript, critically reviewed and edited the manuscript and tables, and approved the final version for publication.

CW designed software programs in R to format and population weight the data, aided with the SAS statistical analysis, critically reviewed the manuscript, and approved the final version for publication.

### Additional Contributors

Martin Sebera, from the Department of Kinesiology, Faculty of Sports Studies Masaryk University, Czech Republic, critiqued statistical aspects of the manuscript and provided useful input. Pavel Grasgruber, from Masaryk University, Czech Republic, provided suggestions after reviewing the manuscript. We thank Scott Glenn and Brent Bell from IHME who supplied us with the GBD risk factor exposure data for the risk factors and for BMI data.

### Data sharing statement

The raw, unformatted data used in this analysis is now out of date. The 2019 GBD data on all the variables in this analysis may be obtained from the IHME by volunteer collaborating researchers. The formatted database, SAS codes and Excel spreadsheets on which this analysis is based are posted on the Mendeley data repository: https://data.mendeley.com/datasets/g6b39zxck4/6

### STROBE checklist

This report follows the STrengthening the Reporting of OBservational studies in Epidemiology (STROBE) guidelines for reporting global health estimates.^36^

